# Performing specialized biological tests with task-specific machine learning models

**DOI:** 10.64898/2026.05.10.26352837

**Authors:** Daniel Bertin, Pierre Bongrand, Nathalie Bardin

## Abstract

In view of the outstanding progress of machine learning (ML) and growing cost of health systems, it is a current challenge to incorporate artificial intelligence tools into actual medical practice. Here we explored the feasibility and reliability of using machine learning to perform an important immunological investigation that currently requires experienced biologists: Anti-nuclear cytoplasmic antibodies (ANCAs) are important markers for vasculitis and they may be evidenced by microscopic examination of cells labeled with patients’ sera. The use of a reliable ML classifier to discriminate between positive and negative samples would increase the rapidity and decrease the cost of immunofluorescence-based ANCA detection.

Here, we tested seven well-documented ML algorithms, ranging from simple models such as decision trees to more complex convolutional neural networks involving millions of adjustable parameter. We studied the feasibility and reliability of classifying 1114 serum samples that had been collected for about 3 years and assayed with conventional procedure. We compared four strategies consisting of assaying either whole microscope fields or individual cell images, and natural images or histograms. The following conclusions were obtained: (i) Several different strategies allowed us to build models stable enough to discriminate between positive and negative samples collected during about 27 months, with a comparison to human classification yielding a kappa index of about 0.7, that may be considered as fairly good and intermediate between the performance of junior and senior biologists. (ii) Task-specific ML models combined with theoretical thinking might provide the most rapid and efficient way of developing a reliable test within the framework of a single institution. (iii) In addition, the interpretability of the simplest model provided some theoretical insight into important classification parameters. (iv) An important point and caveat is that the multiplicity and versatility of currently available tools make it an essential requirement to test repeatedly a given model, that must be chosen as simple as possible, to achieve a reliability compatible with medical use.

It is concluded that our study provides a strong incentive to incorporate task-specific ML tools in well defined medical tests, which might reduce the risk of human errors and pave the way to fully automatic procedures.

## 1 Introduction

### 1.1 Specific problems raised by the use of artificial intelligence to perform medical tasks

During the last few years, a rapidly growing number of reports described computer tools denominated as artificial intelligence (AI) or machine learning (ML) models with a capacity to perform medical tasks with a reliability matching human skill (reviewed in ^1^). Prominent examples are the use of deep learning to perform primary diagnosis ^2^ ^3^ ^4^ or medical image analysis ^5^ ^6^. Obviously, the introduction of these methods into actual medical practice might dramatically increase availability and rapidity of health care, with a much lower cost. However, while a number of AI-involving models are already in use ^7^ ^8^ and although AI was reported to outperform humans in tasks such as analyzing electrocardiograms ^9^ or chest X-ray images ^10^, AI methods remain far from being routinely used to replace human operators. An important reason for this situation is that the “real world” performance of AI was often reported to be lower than reported in “proof-of-concept” studies ^11^ ^12^. It is thus warranted to subject projects involving medical AI to rigorous and specific evaluation procedures ^11^ ^13^ ^14^. Interestingly, it was emphasized in recent reviews on image analysis that it is not feasible to test all available algorithms ! ^15^ ^16^. Here we shall focus on medical diagnosis and particularly on image classification ^15^. Five AI-specific problems may be emphasized:

1. The enormous **versatility** of AI may be viewed as both an advantage and a drawback. An AI model may be viewed as some kind of “formula” that will be applied to the parameters (or “features”) of a dataset sample to derive a prediction. This “formula” includes a high number of fitted (or “trainable”) parameters (sometimes hundreds of thousand or even hundreds of billions of them ^17^) that are fitted by an algorithm allowing to optimize the predictions done on a subset of the starting dataset denominated as the *training* set. Clearly, if the number of trainable parameters is higher than the training dataset size it may be expected to build a model yielding excellent prediction, but the biological relevance of features used for prediction may be low, and the model will yield poor capacity to generated accurate predictions on a *test* dataset different from the *training* dataset (a property usually denominated as **generalizability**). This situation is defined as **overfitting** and represents a major limitation of AI, namely the difficulty to ensure that predictions are based on biologically relevant sample parameters rather than accidental features. Thus, since the quality of a model is estimated on the basis of predictions made on *test* data, this may be overestimated by unexpected artifacts such as so-called *data leakage* that may be defined as an illicit sharing of information between training and test data ^18^. This possibility might be responsible for a progressive decrease of the efficiency of a validated algorithm as a consequence of an evolution of medical conditions ^19^ ^20^. In conclusion, an important requirement consists of checking the generalizability of a model with a *validation* dataset that is as independent as possible from the training dataset. Note that generalizability can be improved by feeding models with well chosen parameters and incorporating suitable “penalties” in the training algorithm.
2. An additional difficulty is due to the enormous **diversity** of algorithms that are often compared in a single study. In addition, each algorithm may generate a high number of models by fitting of additional parameters called **hyperparameters**. Further, the building of a model involves an additional variability due to the involvement of many random steps (e.g. initial parameter initialization). Thus, the probability that a given algorithm be successful by mere chance may be fairly high, generating a need to use *higher significance levels* than in a standard binary trial.
3. Another difficulty raised by many ML models is that training may require much larger amounts of data than conventional regression techniques ^21^. As a consequence, the quality and homogeneity of data is much **more difficult to check**. Thus, it may not be feasible to label manually a sufficient number of images to train complex classifiers. This difficulty was well recognized ^22^ and there is a growing tendency to use so-called **foundation models** ^23^ ^24^ that may train themselves autonomously on features such as images that are either unlabeled ^25^ or incompletely labeled (this is called “unsupervised” or “weakly supervised” training). These “general purpose” ^26^ models are then used to “encode” datasets in order to yield “biologically relevant” parameters that will we used to feed classifiers. An important criterion of the quality of a foundation model is its “robustness”, i.e. insensitivity to differences between medical centers, e.g. use of different image preparation procedures ^27^.
4. A essential point is that the validation of a ML model is strongly dependent on the parameters used to assess its efficiency. Here, it may be useful to recall that two different quality criteria must be considered:

Firstly, when a model is being built, trainable parameters are fitted by minimizing a function that may include two kinds of components: A) an estimate of the discrepancy between predicted and expected classification of the validation samples, this is often called a “loss function”, and B) a regularization parameter, adding a penalty that is intended to favor the choice of biologically relevant parameters. It must be emphasized that convenient default values of these functions are usually included in the classes provided by platforms such as scikit-learn (http://www.scikit-learn.org) or tensorflow (https://www.tensorflow.org). Importantly, this choice may influence the occurrence of phenomena such as “hallucinations” that may be defined as false but plausible outputs of AI models. Hallucinations were reported to be decreased or eliminated by a specific training ^14^. Another example is the reported incapacity of AI models to indicate that they are unable to find a suitable answer, a problem that may be alleviated by a suitable training ^28^.

Secondly, numerous indices were used to account for the quality of AI models ^16^. A detailed discussion of these indices would not be appropriate in this introduction, but the importance of this point deserves being emphasized and this may be illustrated by a simple example: **prediction accuracy**, i.e. the fraction of valid predictions, provides a safe means of validating algorithms used to sort balanced samples between a high number of classes. A well-known example is the discrimination between ten handwritten digits as provided by the MNIST dataset ^29^ ^30^. However, prediction accuracy is not suitable to assess the capacity of a model to perform a binary choice, e.g. to detect a small fraction of positive samples in patient’s sera subjected to routine tests. Indeed, a model predicting that all samples are negative in a population with 1% positive subjects will display 99% prediction accuracy, which might be considered as excellent ! Thus, it is important to remember that the choice of quality criteria is an important requirement to the production of models reliable enough to allow medical use ^31^.

v) In addition to the aforementioned problems, the use of too complex models may result in other unwanted consequences: thus, it was emphasized that academic users might lack the computer power required to use too complex models ^32^, and that the use of these models resulted in an energy consumption that might negatively impact our environment ^17^.

### 1.2 Purpose of this paper

The purpose of this paper was to assess the capacity of different task-specific algorithms to detect autoantibodies associated with autoimmune diseases by analyzing microscopic fluorescence images, as described in several recent reviews ^33,34^. We extended a preliminary report ^35^ with a 20-fold larger dataset. The general principle consists of incubating standard cell samples with patients’ sera, then labeling bound antibodies with fluorescent anti-immunoglobulin antibodies. This procedure may be used to detect the binding of anti-nuclear antibodies (ANAs) to microscopic slides coated with Hep2 cells (a tumor cell line). ANAs are associated with a number of autoimmune diseases such as systemic lupus erythematosus ^36^. In other situations, microscope slides may be coated with blood leukocytes to detect anti-neutrophil cytoplasmic antibodies (ANCAs) that were reported to be associated with vasculitis ^37^. The examination of microscopic images is used firstly to detect positive samples leading to further examinations, secondly to obtain some information on antibody specificity by analyzing fluorescence pattern. Microscopic examination has long been considered as a gold standard for ANA and ANCA analysis ^38^, but this must be performed by an experienced biologist ^39^, which raises the cost of the procedure and increases the waiting time for the conclusion. Thus, immunofluorescence microscopy is more and more often replaced with assays of specific antibodies in patients’s sera with immunochemical methods ^40^ ^41^ ^42^. However, immuofluorescence may still be useful to confirm immunochemical tests, since it may increase specificity for weakly positive samples and sensitivity ^43^. Indeed, currently available immunochemical tests will not reveal new auto-antibody specificities. It might thus be rewarding to perform an automated analysis of immunofluorescence images as a rapid and affordable means of detecting sera that might benefit from a more extensive set of detailed tests.

Here we compared the efficiency of several different methods to analyze 1114 sera collected during a period of nearly 3 years in a hospital immunology laboratory as an extension of a preliminary study made on 137 samples ^44^. The following strategies were studied: use of hand-crafted parameters, direct analysis of whole field images with AI algorithms of varying complexity, image segmentation and analysis of individual images.

We used as a quality reporter the *kappa index* ^45^ calculated by comparing the model prediction and conventional classification as performed by experienced human biologists. The interest of this index is that is is widely used in both medical literature and studies focused on artificial intelligence. It provides a comparison between the efficiency of a classification algorithm and random sorting, yielding values usually ranging between 0, corresponding to random sorting, and 1, corresponding to perfect classification. A negative value is indicative of a classification algorithm that is worse than mere chance. As a rule of thumb, it was suggested that the efficiency of a method might be considered as moderate, substantial or nearly perfect when kappa is respectively 0.4, 0.6 or 0.8 ^45^.

In addition, it was found important to define a reporter of overfitting that may be considered as a major limitation of most machine learning models ^46^, and may be defined as a model inability to give predictions on a test dataset as efficiently as predictions yielded for the training dataset. Thus, we defined an overfitting index as (kappa_train_ - kappa_test_)/kappa_train_. This was expected to range between 0, corresponding to an equal efficiency of the model to classify the training and test datasets, and 1, corresponding to a total lack of predictive power on the test dataset. As a rule of thumb, it may be expected that a low kappa train value is indicative of the model inability to fathom the complexity of the classification task, while a high overfitting index suggests that the model classification is not based on the parameters that were felt of interest in a considered task.

It is concluded that a kappa index on the order of 0.7 can be obtained with several easy-to-use task-specific methods. This may be considered as an incentive to include a computer-assisted analysis in standard hospital procedures, which might lead to both increased safety and gathering of more extensive datasets that might foster future progress.

## 2. Results

Ethanol-fixed human neutrophils deposited on glass slides were labeled with DAPI, a nucleic acid stain, and patients’ sera together with fluorescein-labeled anti-immunoglobulin, to reveal autoantibodies. They were then examined with fluorescence microscopy using two excitation wavelengths to reveal both DAPI and fluorescein. DAPI labeling was used to determine individual cell contours. Representative individual cell images are shown on Figure 1. A total of 1114 serum samples were collected during a three-year period of time and examined by experienced biologists for ANCA detection: 897 samples were classified as negative, and 217 were classified as positive. Image segmentation yielded 36163 individual cell images (29060 from negative samples, 7103 from positive ones).

**Figure 1.**
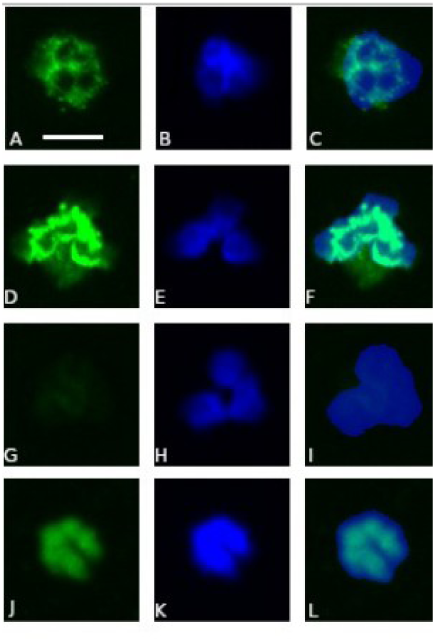
Representative immunoflorescence images of cells labeled for ANCA detection. Cells classified as c-Anca (A, B, C), p-Anca (D, E, F), negative (G, H,I) or other (essentially positive for anti-nuclear antibodies: J, K,L) are shown. On the first column (A, D, G, I), fluorescent antibodies appear as green. On the second column (B, E, H, K), DAPI labeling (mostly nuclei, but also the whole cell area) appear as blue. On the third column (C, F, I, L) DAPI and Fluorescein images are superimposed. Horizontal bar lengh: 5 µm.

### 2.1 A computer-assisted method based on a single hand-crafted index matched human analysis with a kappa index of 0.68 and remained stable for a 3-year period of time

First, 10 series of sera (514 samples) were examined by two experienced biologists and kappa index was calculated to assess the match between their conclusions. Calculated values ranged between 0.507 and 1 (mean 0.84+/- 0.15 SD).

Secondly, as a reference for assessing the efficacy of currently used machine learning tools, we first calculated the discriminatory power of a simple hand-crafted index (denominated as ICARE) developed in our laboratory for automatic detection of anti-nuclear antibodies with immunofluorescence ^47^. The basic principle consisted of comparing the mean fluorescein label of DAPI-positive areas to the fluorescein label of DAPI-negative regions. A sample was considered as positive when ICARE index was higher than a threshold determined in order to match human evaluation. Our series of 1114 samples was randomly divided ten times into a training set (75% of samples) and a test set. The threshold was determined on the training set and used to classify the test set. The mean kappa index determined on the test set was 0.68 +/- 0.03 SD. The kappa index obtained on training sets was 0.71 +/- 0.01 SD. Overfitting was 0.047 +/- 0.059.

Note that the **prediction accuracy**, a commonly used reporter of classification efficiency, was respectively 0.91 +/- 0.005 SD and 0.90 +/- 0.013 on training and test datasets.

An essential reporter of the reliability of an AI model is the stability during a period of time that may involve subtle changes of experimental procedures and reagents in routine practice. This point was addressed by dividing the 1114 sample dataset collected during a period of 34 months into four sequential groups labeled as A, B, C and D. Classification was then performed in each group using (a) the threshold yielding maximum kappa on group A, (b) the optimal threshold determined on the same group. As illustrated on Figure 2, two major conclusions were suggested:

i) optimal threshold values were fairly stable between groups (Figure 2A).
ii) classification performed on groups B, C and D with the threshold parameter determined on group A yielded fairly stable kappa values ranging between 0.68 and 0.8 (Figure 2B).

**Figure 2.**
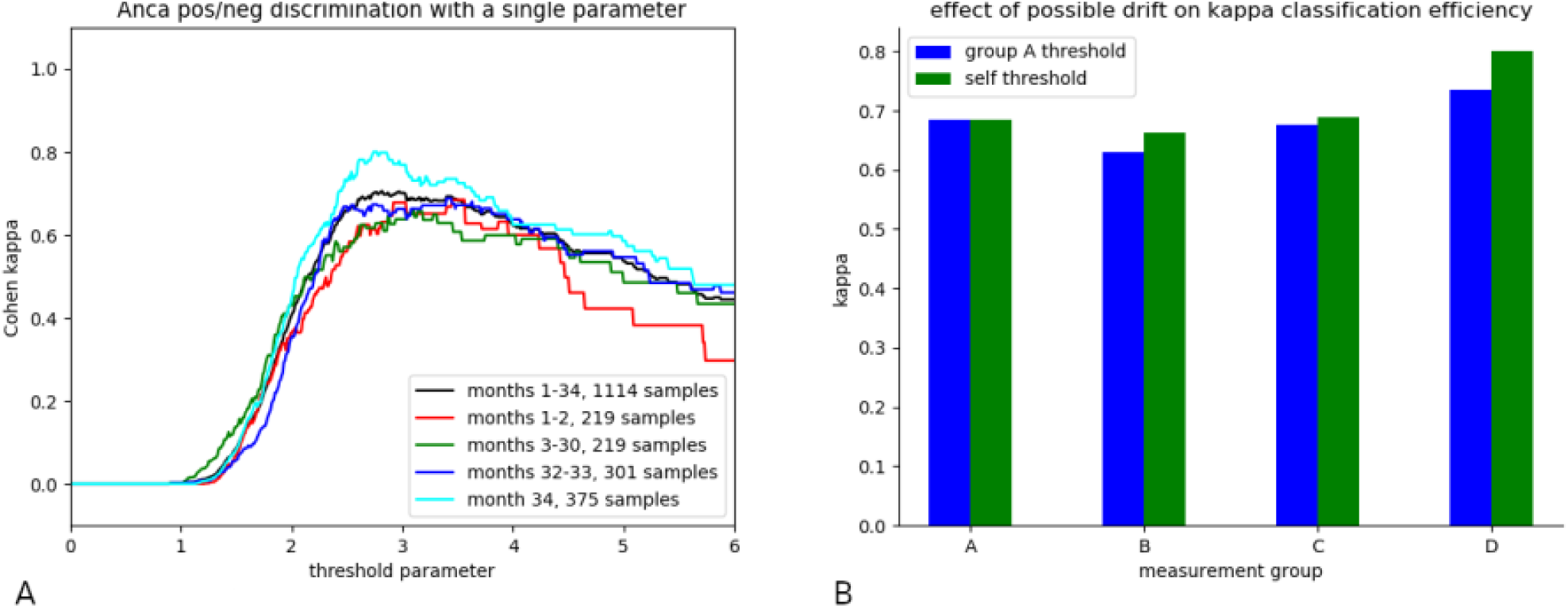
Time stability of Icare classifier. The 1114 images Collected during a 3-year period were divided into four sequential groups (denominated as A, B, C, D). Icare index was determined on all images and a threshold was used to classify images as positive (index higher than a fixed theshold) or negative. Kappa index was calculated to determine classification efficiency. **A - dependence of kappa on threshold parameter**. The threshold value yielding maximum kappa was weakly dependent on sample group and kappa was weakly dependent on the threshold in the neighborhood of the optimal value. **B - Classifier stability**. Kappa parameter was used to evaluate the classification efficiency for all four sample groups, using as a threshold either the optimal value calculated on group A or group optimal value.

Thus a classification model obtained by analyzing the first group could be reliably used during a nearly three-year period of time.

### 2.2 Use of standard machine learning tools to analyze fluorescein label in whole microscope field was less efficient than ICARE

First, we used a number of standard AI methods to analyze fluorescein label in whole microscope fields. As shown on Figure 3, Simpler tools such as nearest neighbor classifier (Knn), decision tree classifier (Dt) and support vector machine (Svm) yielded low kappa values (less than 0.2) with high overfitting and this was not overcome by preprocessing data with scaling or principal component analysis (not shown). The more powerful random forest classifier (Rf) yielded a kappa value of 0.45 +/- 0.03 for test-set classification. Since a perfect match was obtained with training-set classification (kappa=1), and the overfitting index was high (0.55 +/- 0.03), efficiency limitation was due to the model incapacity to discriminate between “relevant” image properties and accidental features of training images.

**Figure 3.**
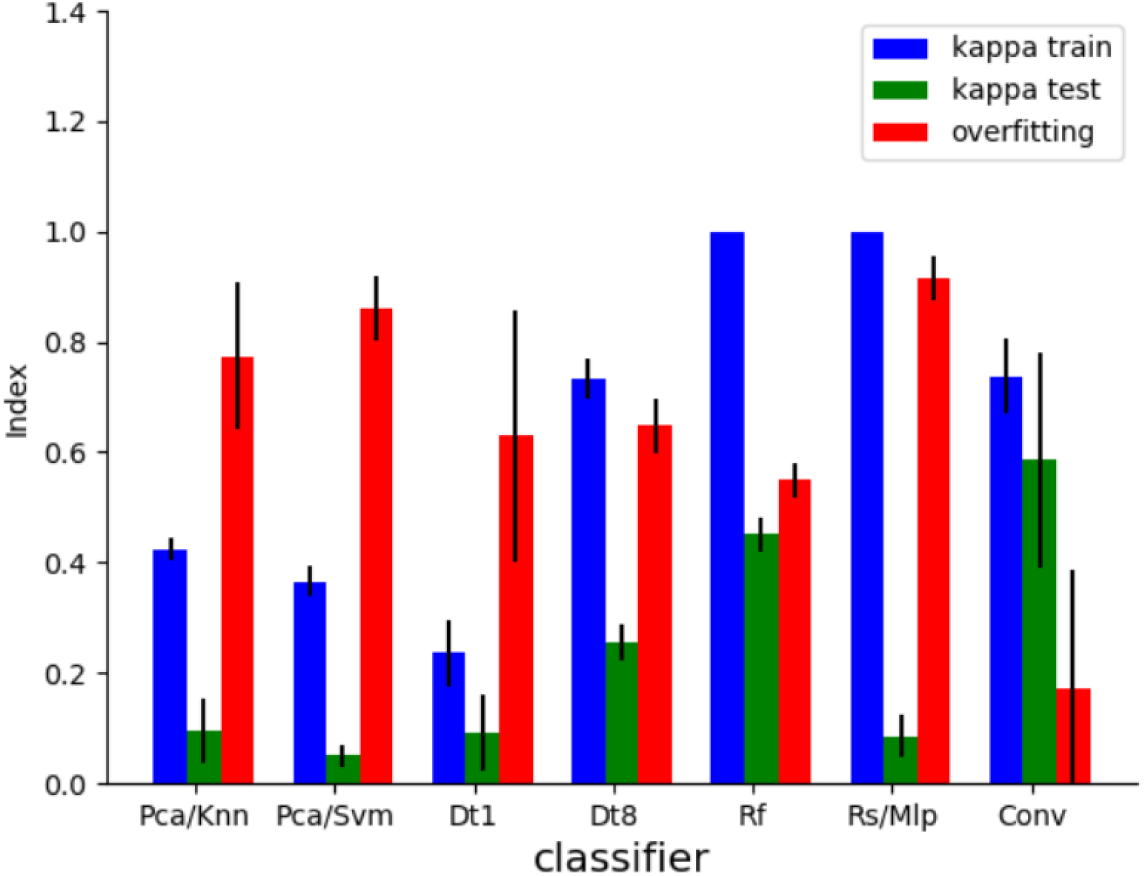
Pos/neg discrimination by processing whole field immunofluorescence images. 1114 whole field fluorescence images were randomly divided five times into a training set (75%) and a test set (25%) for determination of the capacity of seven classifiers to discriminate between positive and negative samples. Mean values of kappa index (calculated for training and test set prediction) and overfitting index are shown. Vertical bar lines are twice standard errors. Classifiers were a) Pca/Knn: K neighbors classifier following dimensional reduction with PCA. b) Pca/Svm: support vector machine following dimensional reduction with Pca. c) Dt1: decision tree classifier with depth=1. d) Dt8: decision tree classifier with depth = 8. e) Rf: random forests. f): Rs/Mlp multilayer perceptron classifier (three layers of 30 neurons each) following scaling. g) Conv: convolutional network (two layers).

Not unexpectedly, the best results were obtained with convolutional neural networks that are known to display powerful image processing capacity. Comparable efficiencies were obtained with networks involving one to three layers (not shown). The kappa index was on the order of 0.6 with an overfitting index on the order of 0.2. However, this value displayed extensive variations with a standard deviation of about 0.2, i.e. between 4-fold and tenfold higher than found with other models.

The stability of these classifiers was also studied: As shown on Figure 4, while the versatility of some AI models allowed excellent match with training datasets, classification of other image series was quite inefficient, suggesting that discrimination of training images was based on features that were irrelevant to the addressed biological problem.

**Figure 4.**
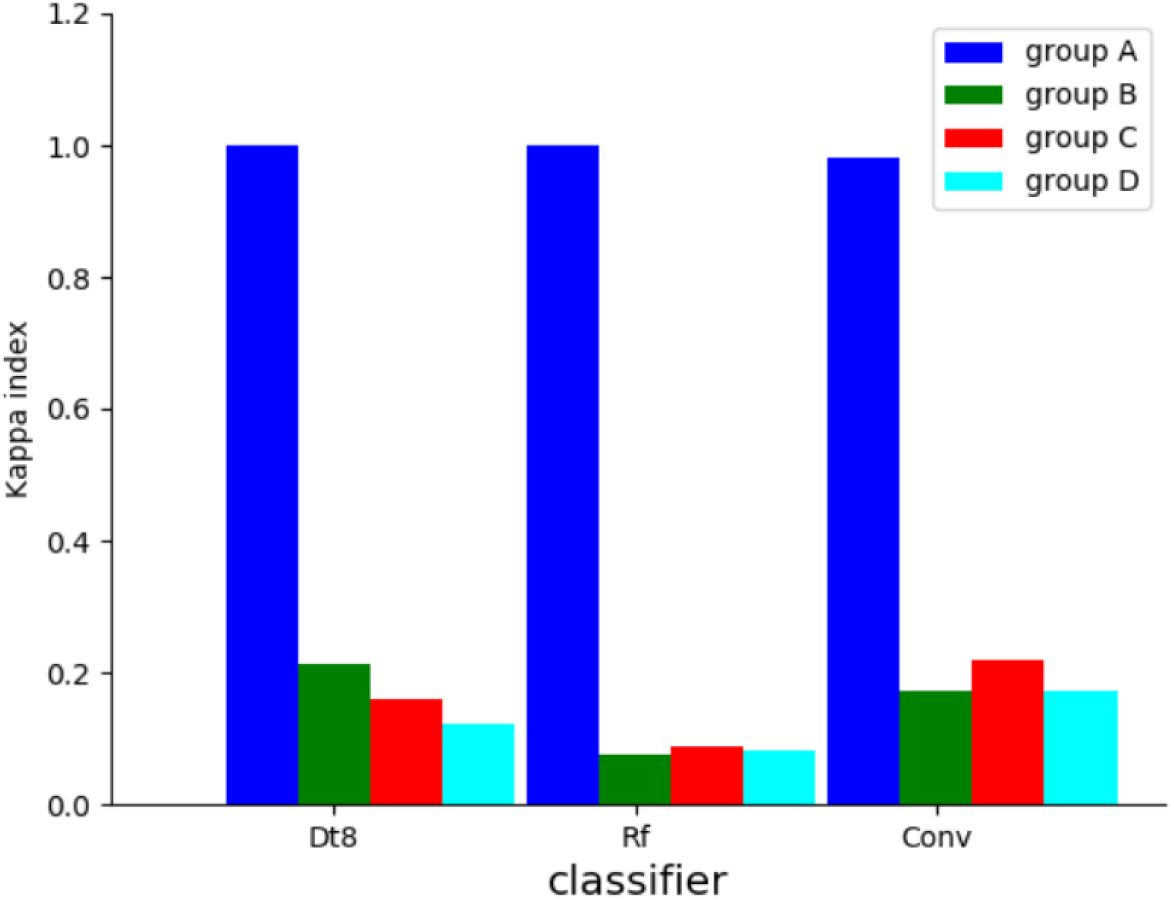
Stability of total image processing. Immunofluorescence images collected during a 3-year period were divided into four sequential groups (labeled A,B,C,D). Three classifiers (Dt8: decision tree/depth=8, Rf: random forests, Conv:2-layer convolutional network) were trained on group A images and used to classifiy the four image groups. Kappa index was calculated and is shown.

Two non-exclusive hypotheses might be considered: i) ICARE superiority might be at least in part dependent on the use (for segmentation) of DAPI images that were not provided to IA models, thus providing ICARE with more information than other models. ii) Due to the high versatility of AI models, they might not focus on features relevant to positive/negative discrimination. The application of these two ideas will now be described.

### 2.3 Replacing standard images with histograms strongly increased ML efficiency

Since ICARE analysis is based on fluorescence intensity distribution within an image, it was reasonable to assume that the information provided by the image histogram should be sufficient to allow a reasonable classification efficiency. Thus, standard classifiers were made to process image histograms (256 intensity levels) rather than whole field images (on the order of 10^6^ pixels). As shown on Figure 5, many classifiers displayed an efficiency comparable to that of ICARE model, with similarly low overfitting index. Unexpectedly, a decision tree classifier with an unusually low depth of one exhibited a kappa of 0.71 +/- 0.05 with an overfitting index of 0.05. The interest of this finding was that this simple model was easily interpretable: classification was performed with a simple thresholding based on the fraction of pixels with a fluorescence intensity near level 60/256.

**Figure 5.**
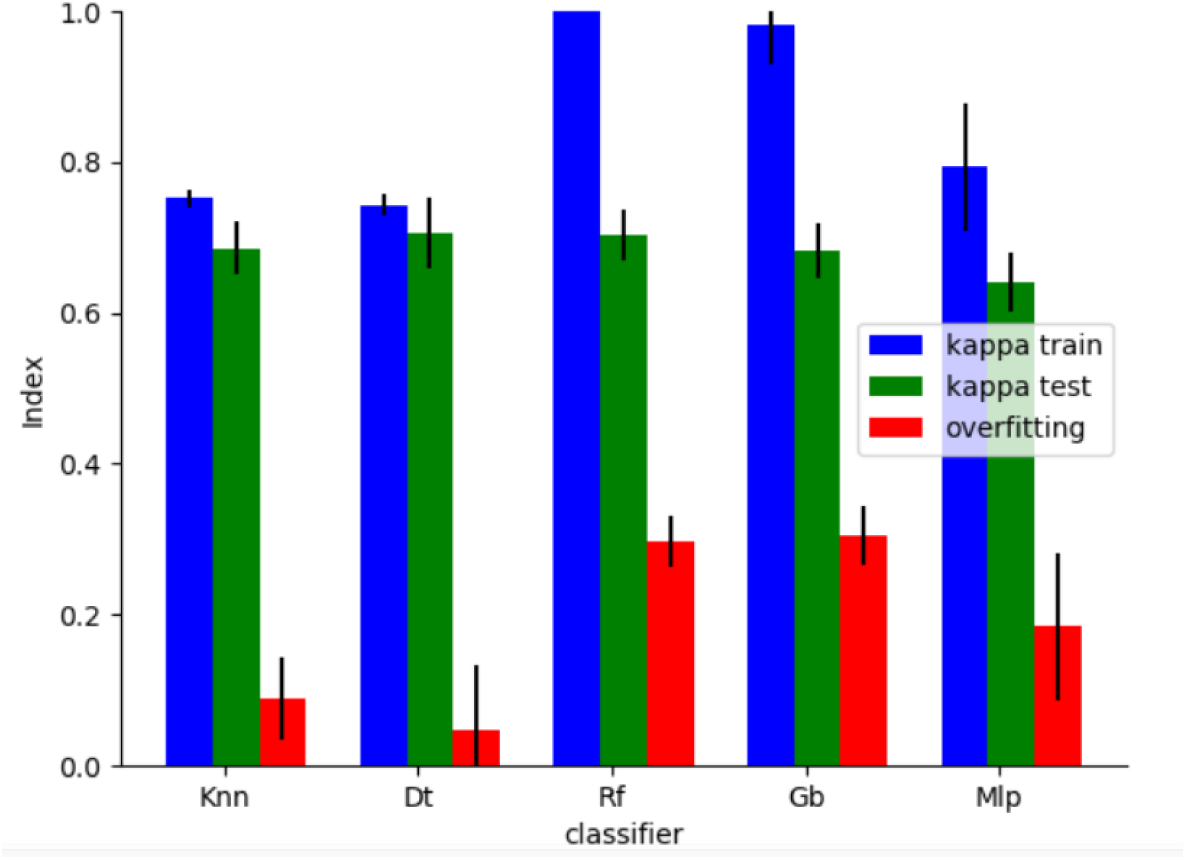
Capacity of several model to derive pos/neg classification from image histogram. The histograms of 1114 whole field fluorescence images were randomly divided five times into a training set (75%) and a test set (25%) for determination of the capacity of five classifiers to discriminated between positive and negative samples. Mean values of kappa index (calculated for training and test set prediction) and overfitting index are shown. vertical bar lines are twice standard errors. Classifiers were a): KneighborsClassifier (Knn), b) Decision tree classifier (Dt, Depth=1), c) Random forests (Rf), d) Gradient boosting classifier (Gb), e) multilayer perceptron classifier (three 30-neuron layers).

The durability of simple models was also studied: as shown on Figure 6, when data were divided into four sequential groups, a model trained on the first group displayed comparable classification efficiency on the following three groups, suggesting that a trained model might be used during several years without any correction.

**Figure 6.**
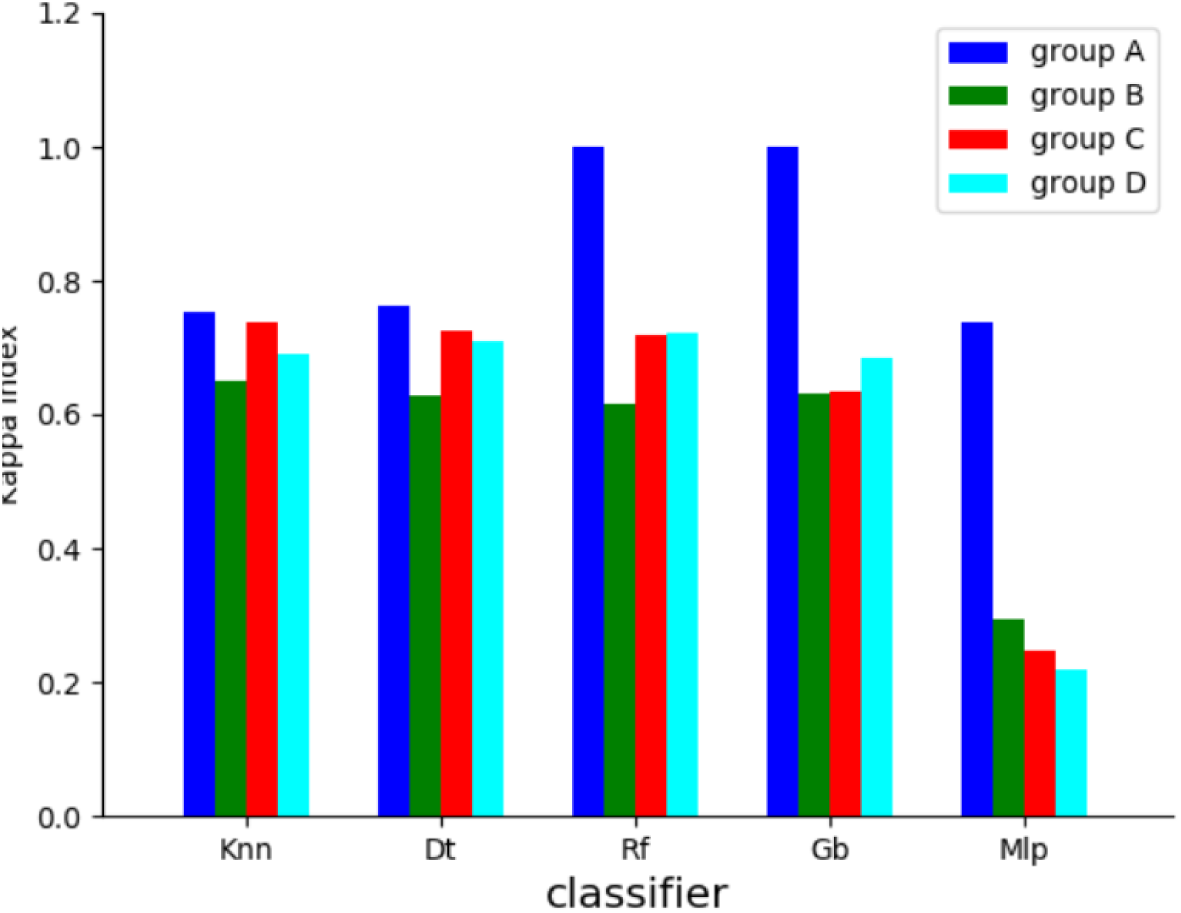
Stability of pos/neg classification with image histogram. The histograms of 1114 immunofluorescence images collected during a 3-year period were divided into four sequential groups (labeled A,B,C,D). Five classifiers (KNeighborsClassifier (Knn), Decision tree (Dt, depth=1), Random forest (Rf), Gradient boosting classifier (Gb) and Multilayer perceptron classifier (Mlp, three 30-neuron layers) were trained on group A images and used to classify the four image groups. Kappa index was calculated and is shown for all four groups.

The following conclusions were suggested by our results:

i) standard ML tools were less efficient than a model based on a single hand-crafted parameter to achieve robust classification of complex datasets when the number of training samples was about 1,000 fold lower than the number of parameters (or features) of each sample. This is in line with the well-known and fairly intuitive principle that a minimal versatility is required for a model to fit a complex dataset, but a sufficient number of samples is required to rule out overfitting as a consequence of the use of irrelevant features by insufficiently trained models. The importance of a correct match between sample and model complexity is illustrated on Figure 7.
ii) Replacing whole images with histograms seemed a convenient means of achieving a substantial dimensionality reduction without any important information loss, since it was reasonable to hypothesize that the image histogram retained properties that were essential for positive/negative discrimination. Interestingly, associating this theoretically inspired procedure with ML resulted in a marked improvement of image classification efficiency as compared to ICARE method, since a satisfactory kappa index could be obtained *without* the use of DAPI labeling, that was required to calculate ICARE index. In addition, the interpretability of the decision tree classifier model was of theoretical interest. This emphasized the interest of combining machine learning with theoretical guidance.

**Figure 7.**
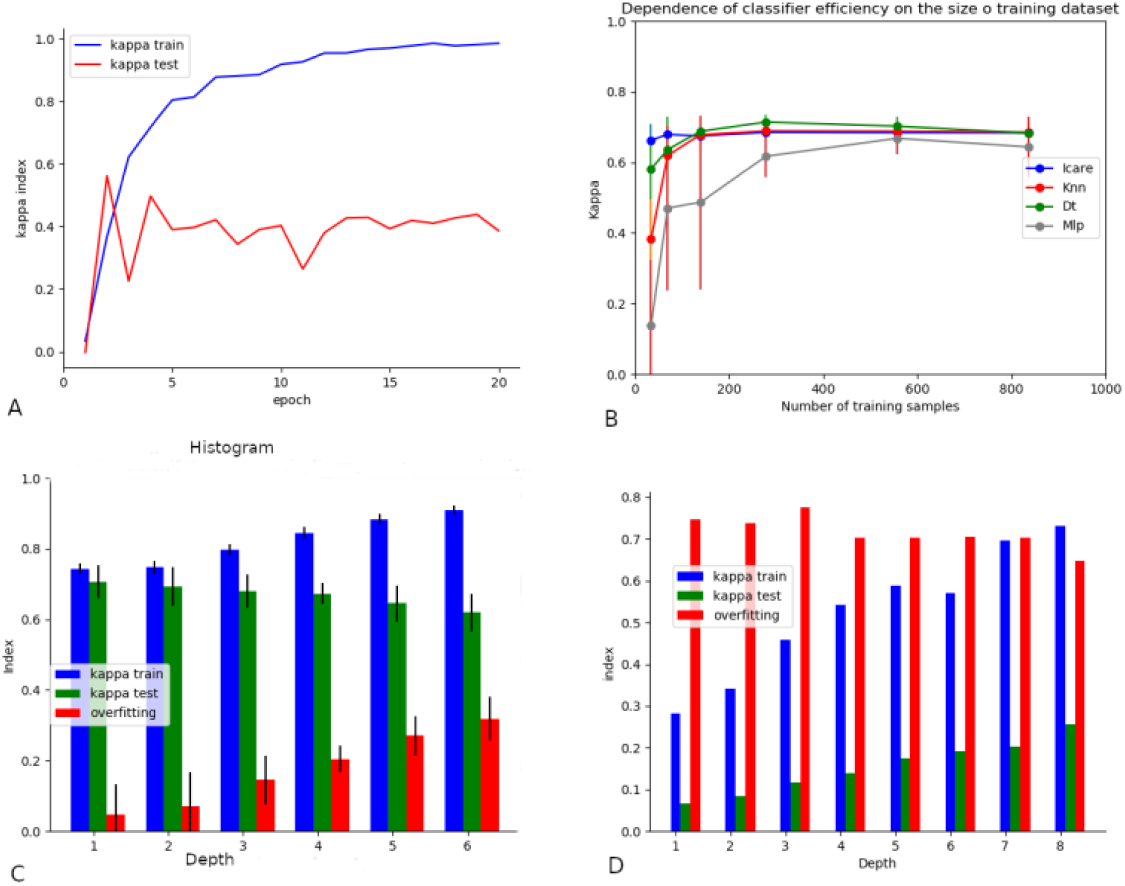
Influence of model and data complexity on classification efficiency. 1114 fluorescence images were used to explore the dependence of classification efficiency on several parameters. Data were randomly split into a training set (75%) and a test set (25%). A) A convolutional network (one layer) was trained for 20 epochs and the evolution of the kappa index of training set and test set predictions are shown. B) The training set was randomly reduced and the dependence of the efficiency of four classifiers on the number of training samples is shown. C) Decision tree classifiers of varying depth (from 1 to 6) were trained on image histograms and efficiency parameters (kappa index for training and test prediction, and overfitting index) were calculated. Mean values of 5 independent experiments are shown. Length of vertical bar length is twice the standard error. Classification efficiency was weakly dependent on classifier depth, with a maximum value for the shallowest classifier. D) Decision tree classifiers of varying depth (from 1 to 8) were trained on whole images and efficiency parameters are shown. The efficiency of train and test classification increased when classifier depth was increased.

These conclusions were an incentive to explore another strategy for image classifications: as suggested by previous attempts at immunofluorescence image classification ^33^, analysing individual cell images was expected to be much easier than processing microscopic fields including nearly 1,000 times more pixels.

### 2.4 Attempts at individual image classification

#### 2.4.1 50×50 pixel single cell images

The 1114 whole field images used above were subjected to DAPI-based segmentation, yielding a total of 36363 single cell images. The human based classification of these fields was used to label individual cells, and classifiers were subjected to supervised training. Representative results are displayed on Figure 8. The following conclusions were obtained:

i) All tested classifiers yielded fairly satisfactory efficacy, since kappa test ranged between about 0.59 and 0.69, which was a marked improvement as compared to results displayed on Figure 3.
ii) As expected, increasing the number of training samples and decreasing their complexity resulted in marked decrease of overfitting.
iii) Interestingly, Random forests (Rf) and Multilayer perceptron (Mlp) differed from simpler models such as Knn and Svm with higher capacity to fit training samples, higher overfitting and comparable efficiency on test sets.
iv) It must be noticed that the low overfitting of convolutional models could be ascribed to a fairly different training process: Rf and Mlp were fitted with fully automatic convergence, as performed with scikit-learn fit function. In contrast, the training plot yielded by convolutional model training (as exemplified on Figure 7A) was used to chose the parameters yielding optimal test set classification, which were obtained after a few epochs. This procedure, denominated as early stopping, is considered to reduce overfitting.
v) The similarity between the efficacy of highly different classifiers is consistent with the hypothesis that all models were able to perform a task as simple as positive/negative discrimination and extract similar basic pieces of information with similar criteria.

**Figure 8.**
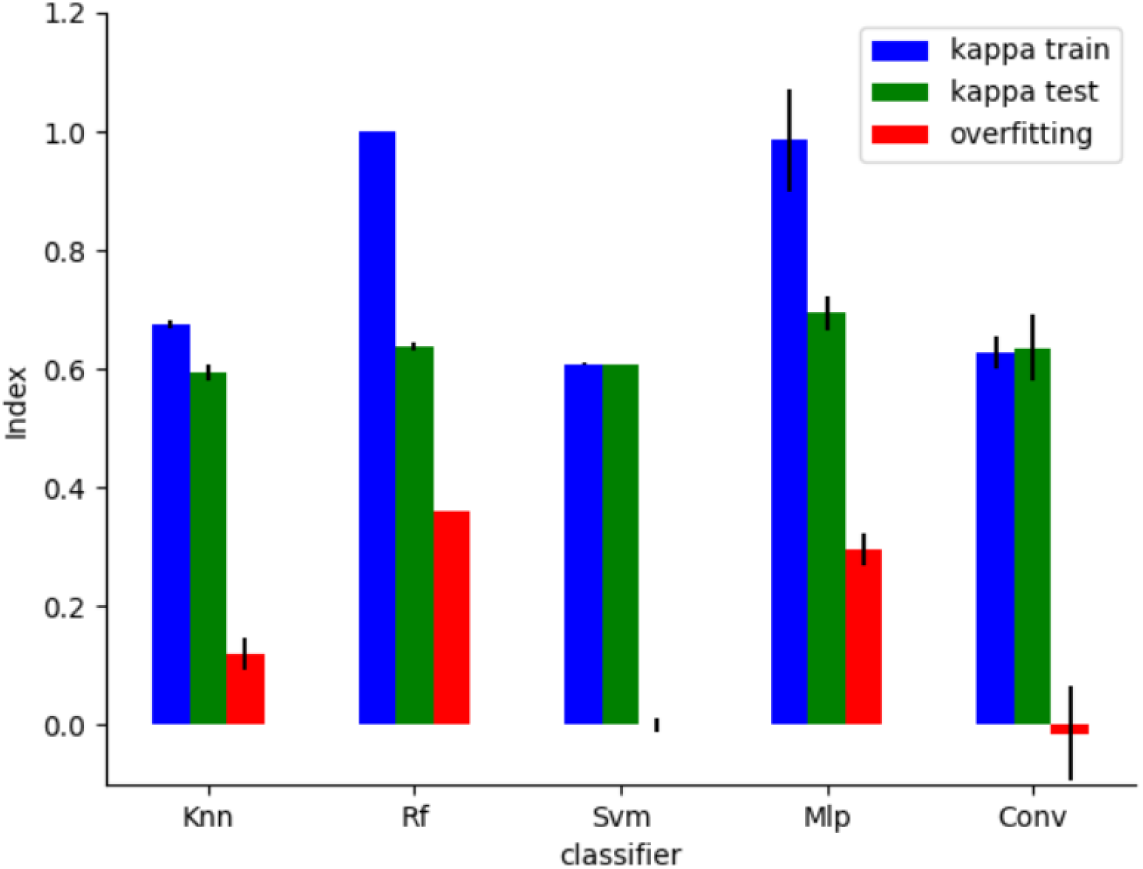
Efficacy of individual cell image classification. DAPI labeling was used for segmentation of 1114 whole field images, yielding 36163 50×50 pixel images containing individual cells. Five classifiers (Knn: nearest neighbors classifier, R: random forests, Svm: support vector machine, Mlp: multilayer perceptron classifier, Conv: convolutional network) were used for supervised positive/negative classification. The image dataset was randomly divided five times into a training set (75%) and a test set (25%). The mean kappa index calculated on the training and kappa set, and overfitting index are shown. Vertical bar length is twice the standard deviation.

We wondered whether classification performance could be still improved by simplifying classifier task and replacing full cell images with histograms, since these histograms were hypothesized to include essentially all information relevant to positive/negative discrimination.

#### 2.4.2 Histograms of single cell images

As shown on Figure 9, replacing cell images with histograms resulted in marked decrease of overfitting index and a possible (not highly significant) improvement of classification efficiency. Thus, after performing five random splittings of training and test data sets, the kappa index of test dataset classification increased from 0.595 +/- 0.013 SD to 0.705 +/- 0.0348 with knn, and from 0.694 +/-0.029 SD to 0.711 +/- 0.024 SD with Mlp.

**Figure 9.**
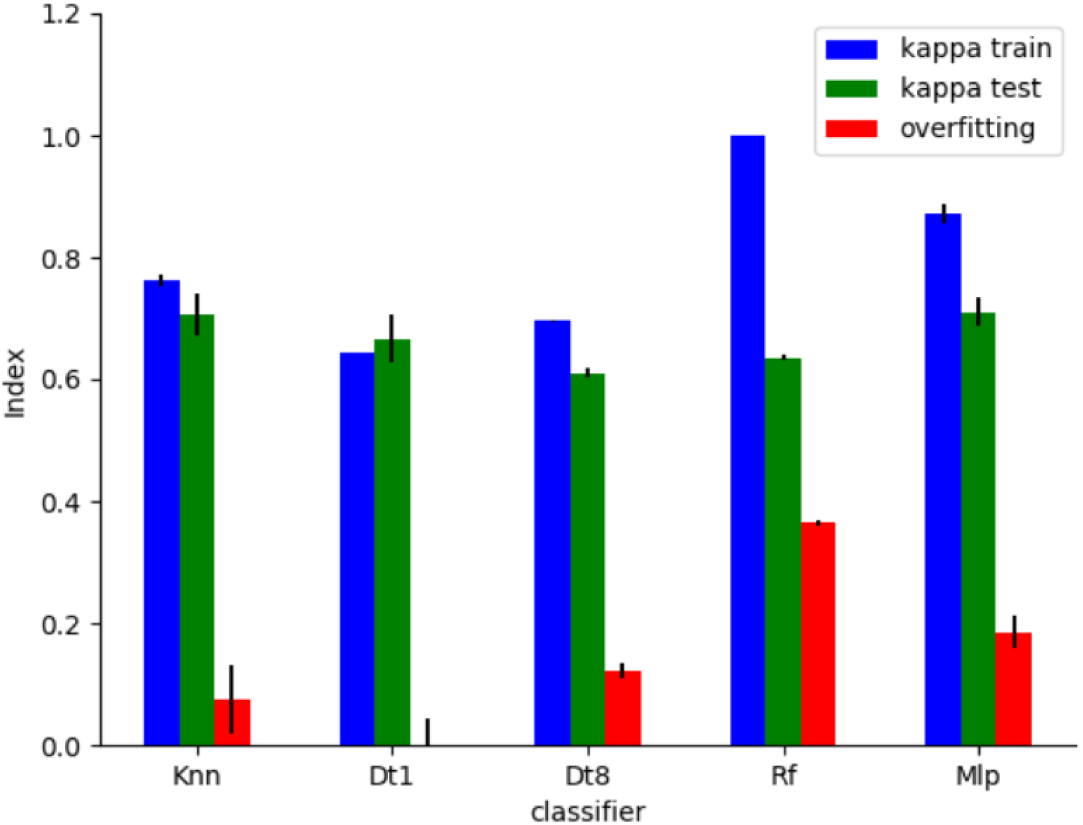
Efficacy of individual cell image histogram classification. The histograms of 36163 50×50 pixel images containing individual cells were used. Five classifiers (Knn: nearest neighbors classifier, R: random forests, Svm: support vector machine, Mlp: multilayer perceptron classifier, Conv: convolutional network) were tested for supervised positive/negative classification. The image dataset was randomly divided five times into a training set (75%) and a test set (25%). The mean kappa index calculated on the training and kappa set, and overfitting index are shown. Vertical bar length is twice the standard deviation.

#### 2.4.3 Tentative improvement of dataset classification with a voting process

The main difficulty encountered in sample classification by human biologists is likely due to the heterogeneity of cell labeling, and this might hamper ML classification. Since the purpose of image analysis is to classify sera rather than individual cell images, it was reasonable to perform this classification by ascribing to each sample the calculated class of the majority of single cell images. The results obtained with this voting procedure are displayed on Figure 10: Interestingly, the voting procedure increased the classification efficiency of all tested models. Thus, in the eight representative examples displayed on Figure 10, The average kappa of test set classification increased from 0.64 to 0.70.

**Fig 10.**
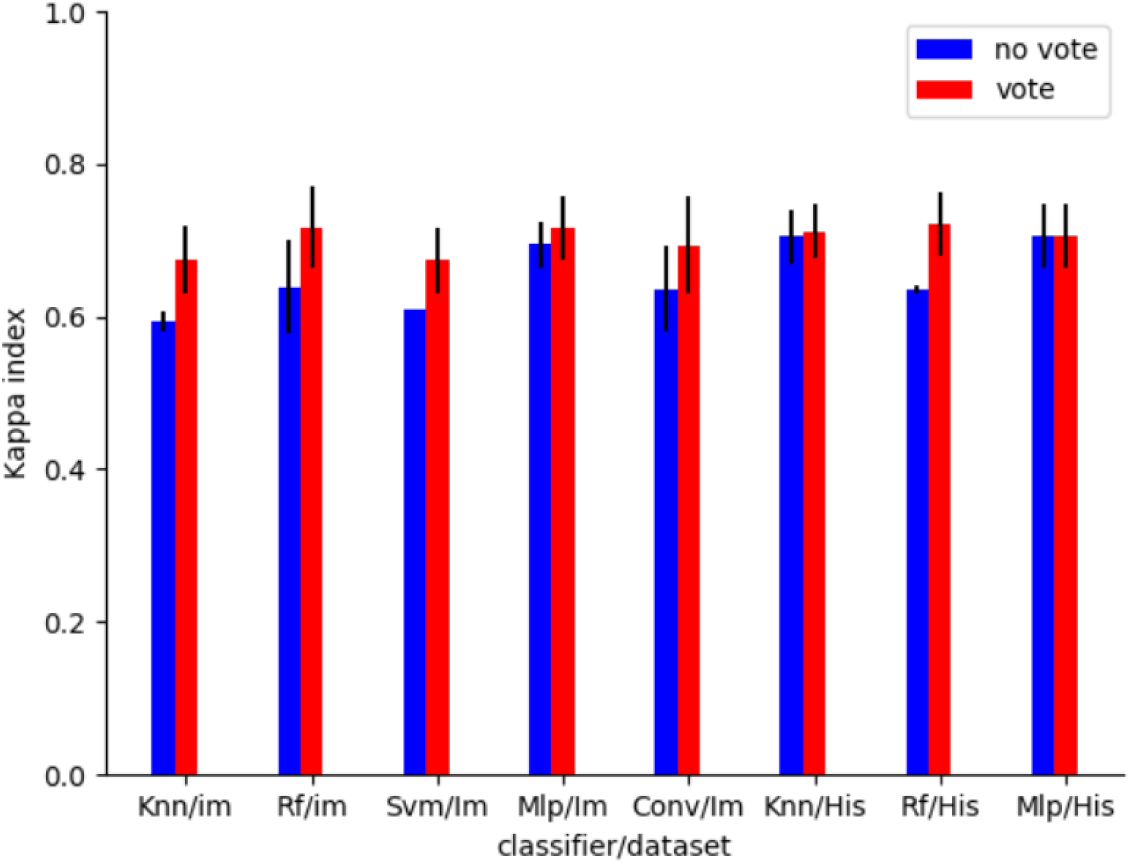
Efficacy of whole field classification with individual cell images. The 36163 50×50 pixel images containing individual cells or 256-level histograms of these images were used. Eight classifier/dataset combinations (Knn: nearest neighbors classifier, Rf: random forests, Svm: support vector machine, Mlp: multilayer perceptron classifier, Conv: convolutional network, im: normal image, His: histogram) were tested for supervised positive/negative classification. The image dataset was randomly divided five times into a training set (75%) and a test set (25%). Classification efficiency was calculated either on individual images (blue bars) or on whole field images that were classified according to the majority of individual cells (red bars). Vertical bar length is twice the standard error.

Since the efficiency of the voting procedure might be related to a possible heterogeneity of cell populations, we asked whether a similar increase of kappa test might be obtained with microscopic detection of antinuclear antibodies, since in this case fluorescence tests are performed with Hep-2 cell line that might be expected to provide more homogeneous cell populations than neutrophils. We addressed this hypothesis by analyzing a dataset of 57018 cell images (20971 negative, 36047 positive) obtained by analyzing 586 serum samples analyzed in our laboratory with a methodology similar to that used for ANCA testing ^47^. We found that the voting procedure induced a similar increase of classification efficiency. Thus when the random forest algorithm was used to analyze these images, the kappa obtained for test set classification was increased from 0.6824 +/- 0.0024 SD to 0.7686 +/- 0.0609 (not shown). However, it must be noticed that Hep-2 cells might also display a marked heterogeneity related to differences in their place in the cell cycle, in contrast with neutrophils.

The influence of voting on the classification stability was also studied. As shown on Figures 11 & 12, voting resulted in a moderate increase of kappa while retaining the general stability of tested classifiers over time.

**Figure 11.**
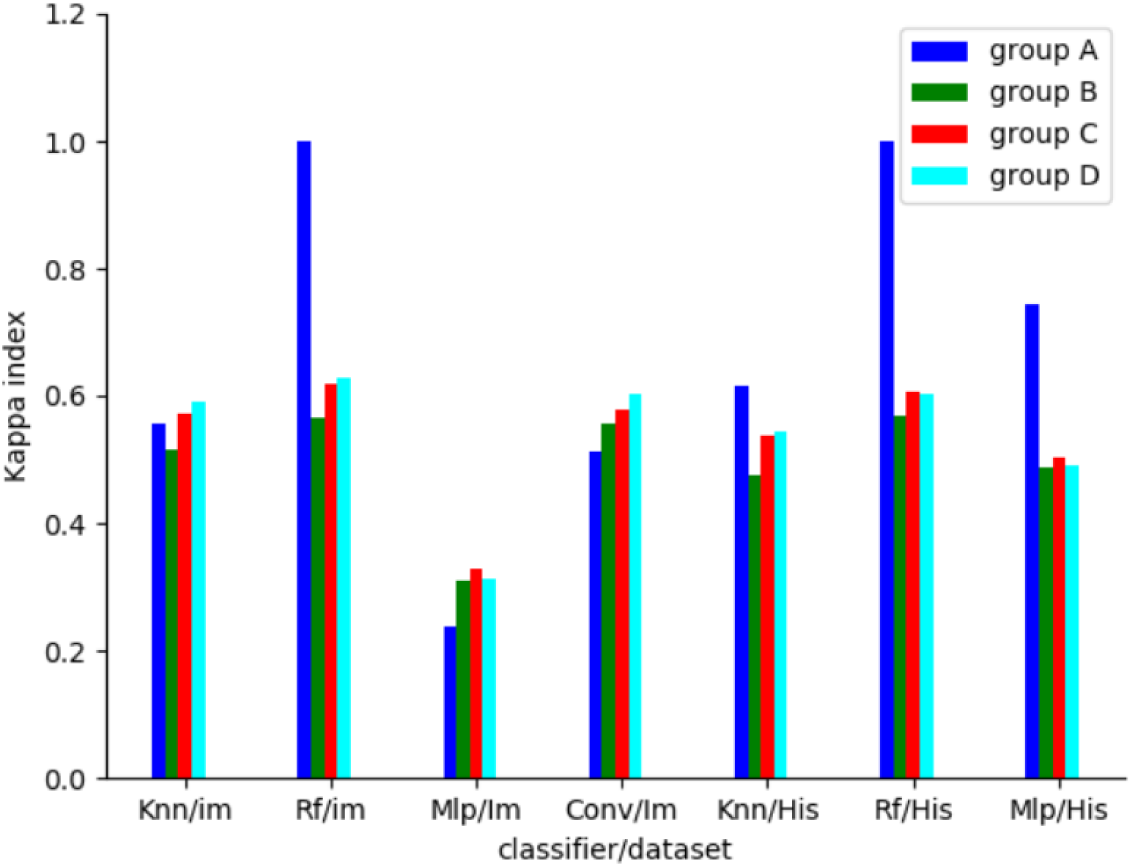
Stability of individual cell image classification. *Immunofluorescence whole field images collected during a 3-*year period were divided into four sequential groups (labeled A,B,C,D). Images were segmented and classifiers were trained on 50×50 pixel images (im) or image histograms (His) of group A. The efficiency of images classification was then determined on all groups and kappa index is shown.

**Fig 12.**
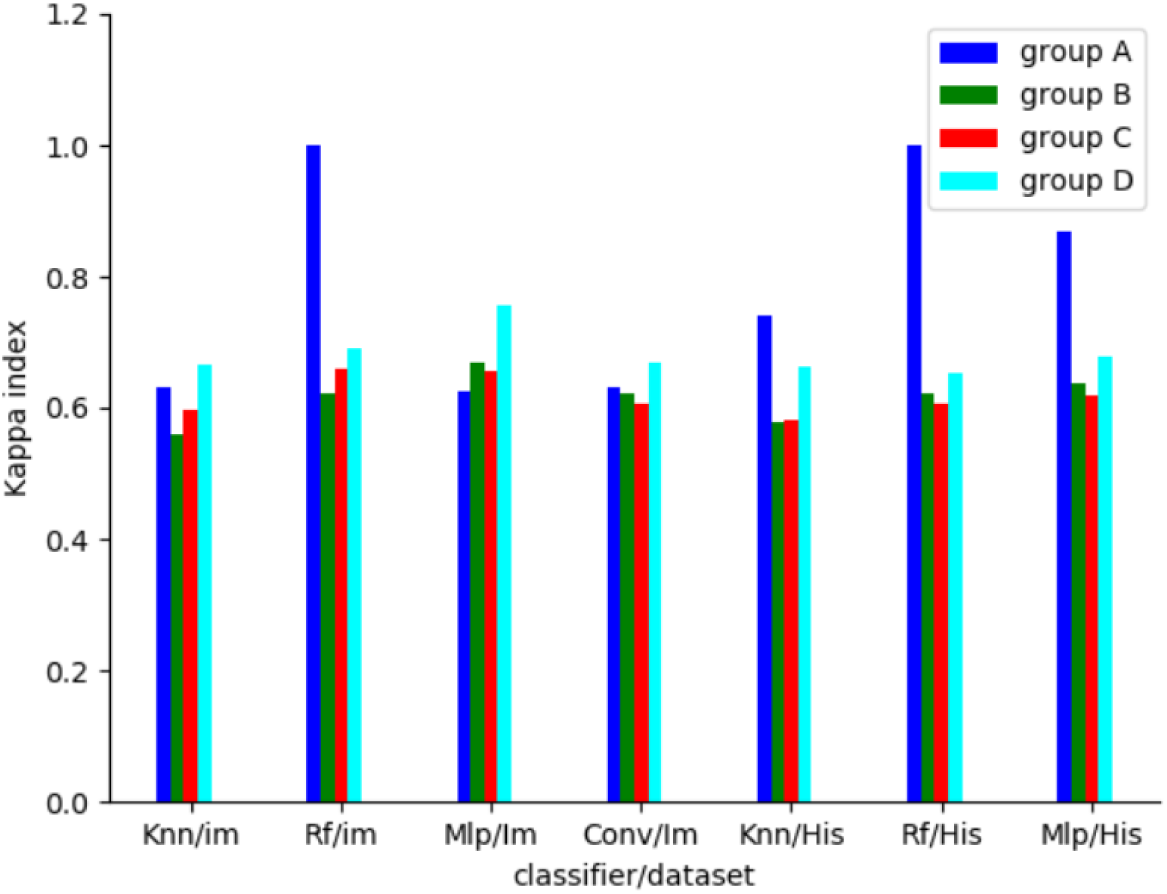
Stability of serum sample classification by processing individual cell images. Whole field immunofluorescence images collected during a 3-year period were divided into four sequential groups (labeled A,B,C,D). Images were segmented and classifiers were trained on 50×50 pixel individual cell images (im) yielded by group A or histograms of these images (His). Whole fields images were then categorized as the most frequent classification of individual images they contained. The efficiency of sample classification was then determined on all groups and kappa index is shown.

### 2.5 A more extensive dataset would be required to compare the efficiency of tested models and strategies

An important aim of this study was to identify the most effective way of using standard ML models for positive/negative discrimination. For this purpose, more than 200 combinations of models and data processing methods were assayed and the frequency distribution of kappa index measured on test samples is shown on Figure 13: Clearly, while a few protocols could be safely discarded since they yielded a very low kappa index, numerous methods yielded a kappa index ranging between about 0.7 and 0.8, and the width of this interval was of the same order of magnitude as the statistical variations of measured kappa indices.

**Figure 13.**
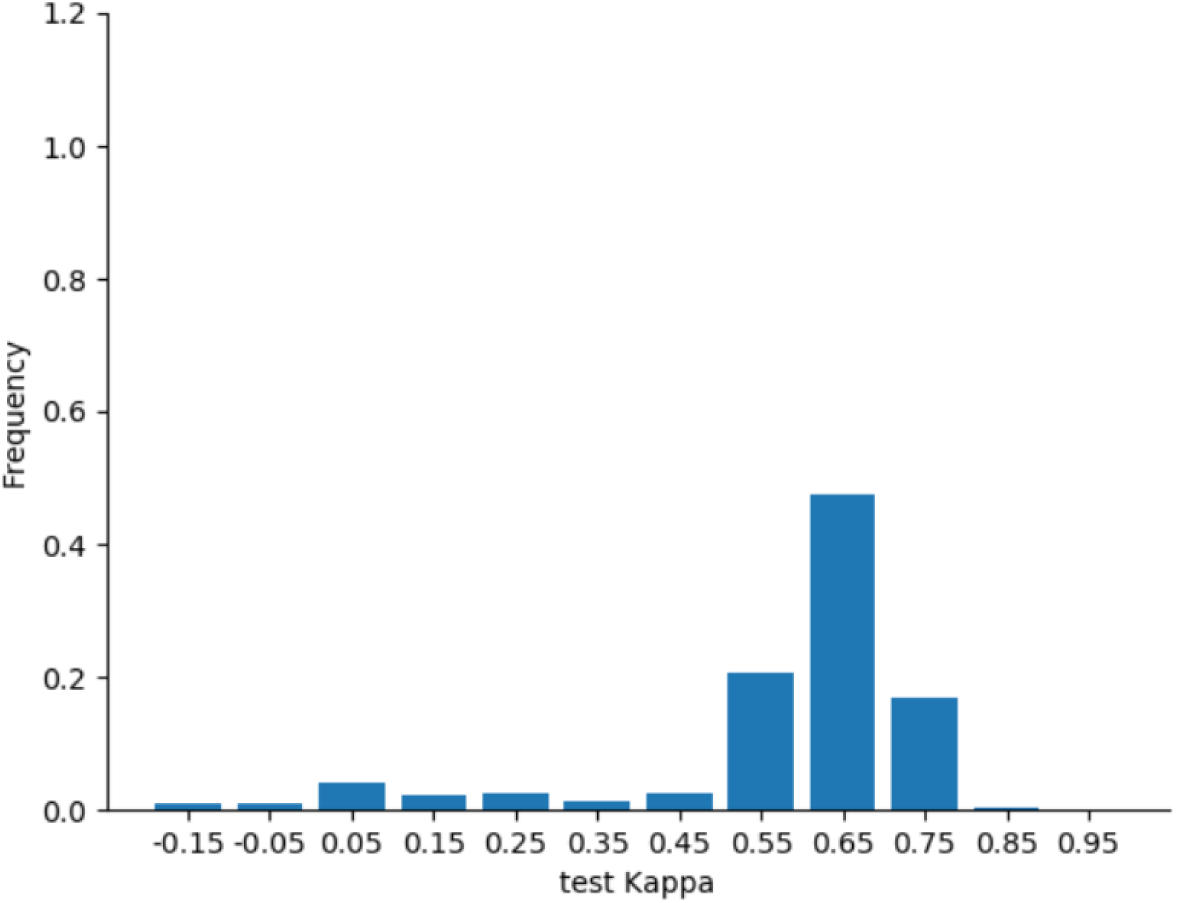
Frequency distribution of test Kappa values. 1114 whole field fluorescence images were processed for positive/negative discrimination with 8 well-documented machine learning models and four data presentation strategies (analysis of whole fields or individual cell images, using actual images or histograms). A total of 229 combinations were considered. In each case, data were randomly split into a training and a test data set. The frequency distribution of kappa index obtained for test set classification is shown.

### 2.6 The main models used in our study readily detected small geometrical patterns

The efficiency of positive/negative classification with histograms showed that the distribution of fluorescence intensities encompassed much information required for classification. It was of interest to determine whether tested models might have also detected geometrical patterns. We addressed this question by using these models to classify artificially modified images where positive samples had been modified by inserting a 6-pixel rectangle of intensity equal to the average background intensity. As shown on Figure 14, this added “clue” significantly improved classifier efficiency, even when a single pattern was added to whole field images.

**Figure 14.**
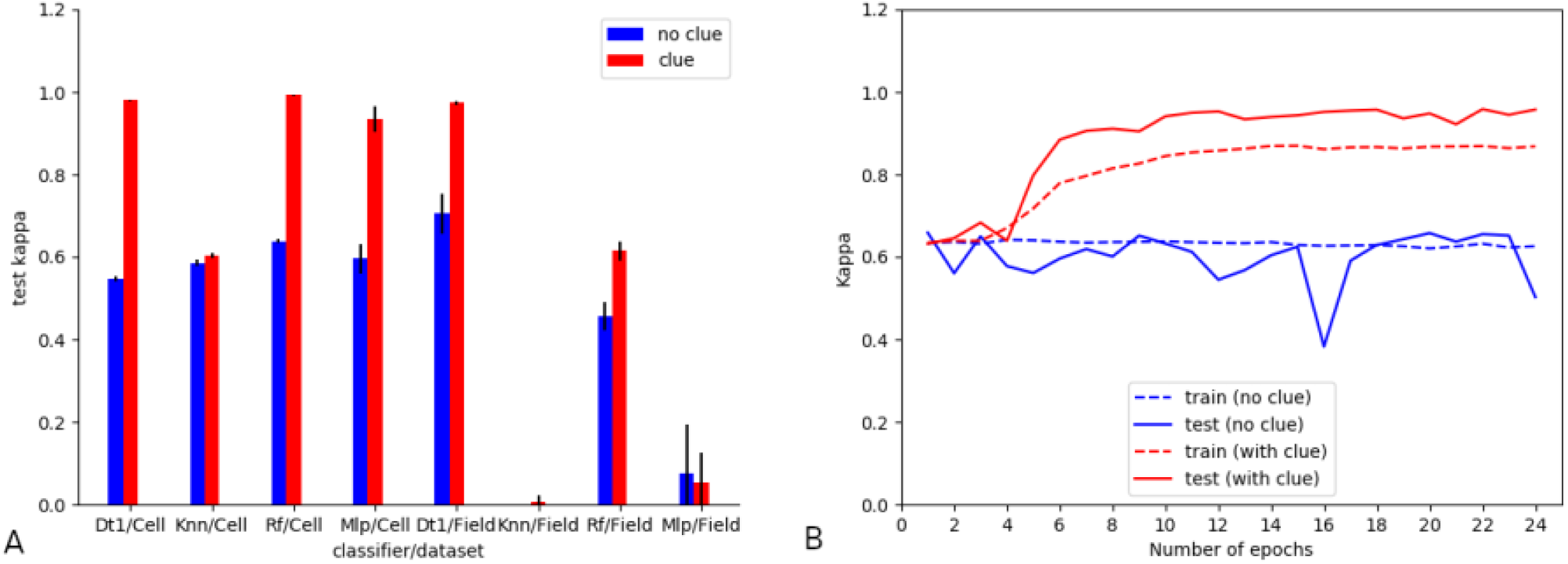
Effect of a simple clue on positive/negative discrimination. A total of 1114 sera were used for immunofluorescence-based detection of Anca. Dapi-based segmentation of 1114 whole field images generated 36163 single-cell images. Machine learning classification was performed (i) on unprocessed cell images and (ii) on datasets where sampled labeled as positive were supplemented with a 3×2 pixel area with uniform intensity close to the background level. A - four models (D1:decision tree with depth=1, knn: k neighbours classifiers with 5 neighbours, Rf: random forests, Mlp: dense neural network with three layers of 30 nodes each. The kappa index of test set classification is shown after processing either single cell (Cell) or whole fields images (Field). Normal images: blue bars. Clue-supplemented images: red bars. Vertical bar length is twice the standard deviation obtained after five independent determinations. B: A convolutional network (2 layers) was trained for supervised classification of the dataset of 36163 normal (blue curves) or clue-supplemented (red curves) single cell images. Images were divided into a training set (75%) and a test set (25%). The kappa set calculated along with model training (training set, broken lines) or after each epoch (test set, full lines) is shown.

As expected, a convolutional network displayed high sensitivity to this pattern since this clue allowed nearly perfect classification after a training of several tens of epochs.

### 2.7 Similar conclusions would have been obtained with different reporters of classification efficiency

While we used kappa index as a single reporter of classification efficiency, it was of interest to know whether the use of other widely used indices would have yielded similar results, since discrepancies between commonly used markers were reported ^48^. This point was addressed by comparing kappa index to prediction accuracy (pa), area under roc curve (auc), and F1-score in 82 randomly selected combinations of models and preprocessing methods. As shown on Figure 15, the three aforementioned indices exhibited close correlation with kappa. As an example, the estimated values of pa, auc and F1 yielded by using a decision tree of depth unity to classify whole field histogram were respectively 0.91, 0.83 and 0.80s as compared to a kappa index of 0.71. Arguably, this might be considered as the simplest procedure. Importantly, it must be emphasized that the relationship between different reporters is dependent on the dataset.

**Figure 15.**
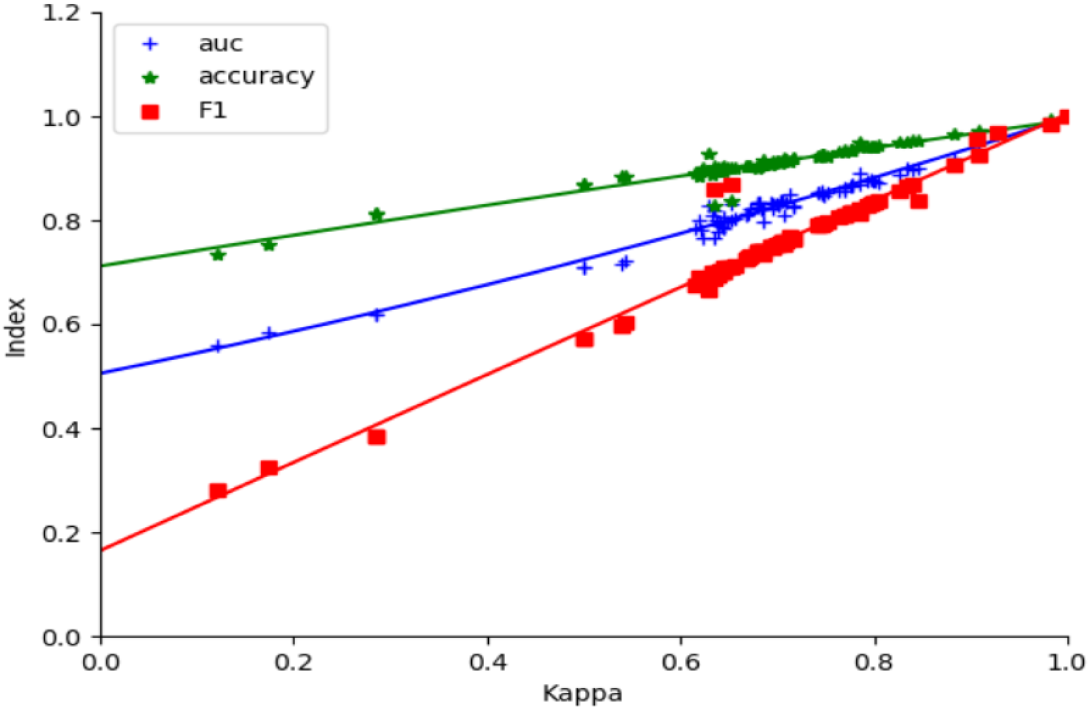
Relationship between several reporters of classifier efficiency. Eighty-two combinations of ML models and preprocessing strategies were used to classify 1114 whole field immunofluorences images for positive/negative discrimination. In each case, kappa index, prediction accuracy, auc (area under roc curve) and F1-score were calculated. The values obtained for the last three indices are plotted versus kappa. Curves represent polynomial fits of degree 2.

### 2.8 Classification was not improved by tentative encoding of images with a classifier trained on a large dataset

The recent interest of general purpose foundation models was an incentive to ask whether pos/net discrimination might be improved by initial encoding of images with a successful classifier. We tentatively used Resnet50, a CNN with 25636712 trainable parameters that was trained on hundred of millions of 224×224 pixel images to discriminate between 1,000 different animal species as defined in Imagenet challenge ^49^. This was used to contribute the analysis of immunofluorescence images ^50^. Interestingly, when cell images were processed and replaced with 1,000-parameter encoding before analysis with different models, a kappa index on the order of 0.7 was obtained with all tested models, as summarized on Figure 16.

**Figure 16.**
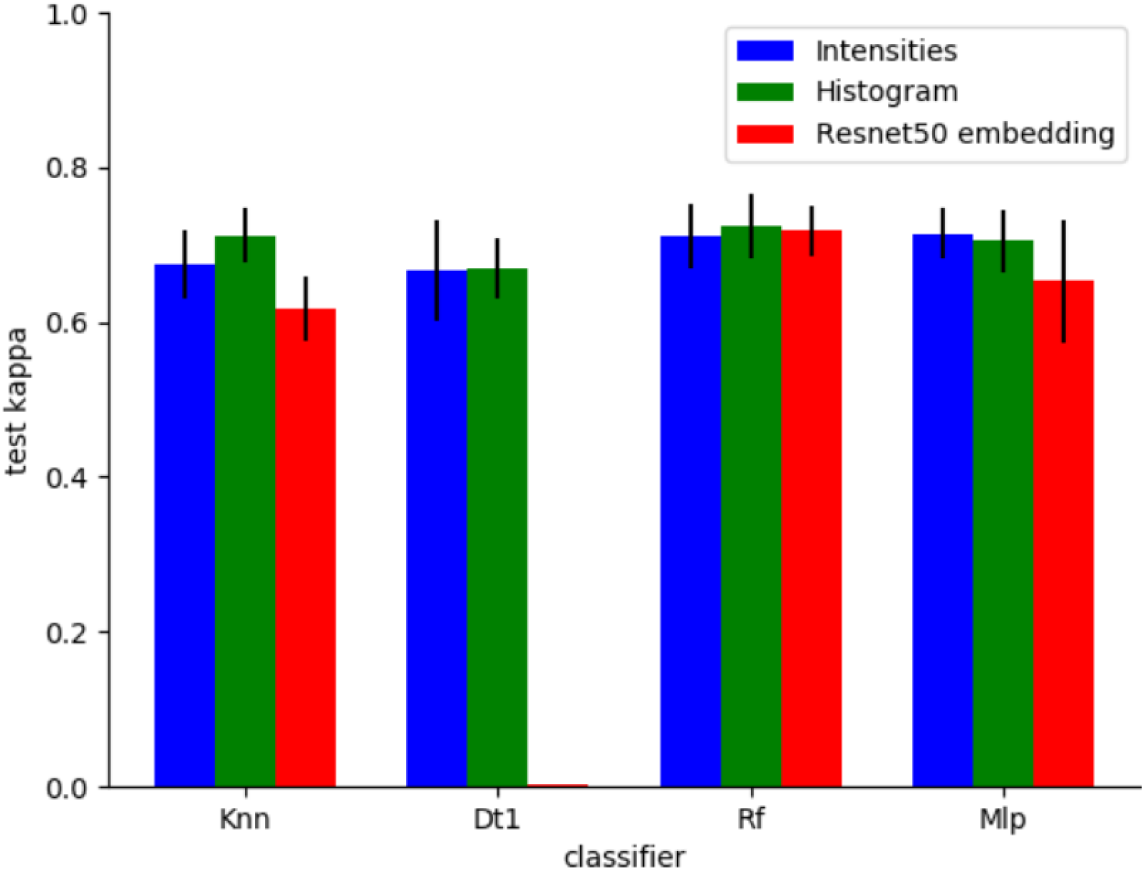
Influence of dataset preprocessing on classifier efficiency. A dataset including 36163 50×50 pixel images of individual cells was used for positive negative classification with four models: Knn (Kneighbors classifier, five neighbors following scaling), Dt1 (Decision tree classifier, max-depth parameter set at 1), Rf (Random forests), MLP (Multilayer perceptron, with 3 layers of 30 nodes each, following scaling). Classifier were trained (i) on image intensities (Blue bars), (ii) 256-level intensity histograms (green bars) or (iii) classification obtained with pretrained Resnet50 (1000 classes; red bars). Kappa value obtained on test classification is shown together with standard deviation (vertical bar lines).

### 2.9 Machine learning models generated similar classification outputs

Since the kappa index yielded by outputs of tested models was subtantially lower than 1, it was interesting to ask whether a voting combination of several classifiers might increase the quality of results. This question was addressed by performing a random splitting of our 1114-image dataset and performing standard analysis with five models: ICARE, decision tree (depth parameter set as one), Kneighbors classifier, multilayer perceptron or a convolutional network: the kappa index obtained with individual models ranged between 0.67 and 0.70, in agreement with aforementioned data, and the voting process only yielded a modest increase of kappa to 0.72. Further, the kappa index obtained by comparing individual models and the voting result ranged between 0.89 and 0.95. Thus, these fairly different algorithms yielded fairly comparable outputs.

## 3. Discussion

The main purpose of this study was to assess the reliability of using task-specific ML models of limited complexity to perform a simple biological task under realistic conditions of hospital practice. A secondary goal was to elaborate simple guidelines to facilitate the choice of a suitable algorithm and training strategy. The following conclusions were suggested:

Firstly, when the performance of tested models was assayed with standard protocols consisting of randomly splitting datasets between a training and a test set, kappa values of 0.7 were repeatedly obtained for test set classification (Figure 13). As a comparison, it was reported that when six powerful pre-trained neural networks were made to classify fluorescence patterns displayed by Hep-2 cells used to detect anti-nuclear antibodies (1,985 samples), kappa values ranged between 0.63 and 0.82 ^39^.

Secondly, as summarized on Figures 6 and 12, our results strongly support the hypothesis that a suitably trained model retained its capacity to classify images collected in an hospital during more than two years with a kappa index higher than 0.6. This may be deemed lower than the performance of an experienced biologist, but higher than that of a junior one that may be on the order of 0.5 as recorded in our laboratory and reported elsewhere ^39^.

Thirdly, an important conclusion is that the high diversity of models and model parameters as well as the intrinsic randomness of the building of many ML models **makes it difficult to identify the best model with reasonable statistical accuracy**. Indeed, a seemingly reasonable aim of our study would have been to identify the “best way” of classifying serum samples. Accordingly, as summarized on Figure 13, it would be tempting to conclude that the strategy yielding the highest test kappa value might be considered as optimal: The highest test kappa score was equal to 0.815, and this was obtained by using a two-layer convolutional network to classify individual cell images and using a voting process to classify serum samples. This is close to the score of 0.84 previously found in an preliminary study made in our laboratory on a sample of 1,733 cell images, i.e. about 20 fold lower that that used in this study ^47^. Interestingly, this best score was also obtained with a voting process. Unfortunately, this seemingly reasonable conclusion may not be considered as tenable: the combination of two-layer convolutional network and voting was performed in 14 independent set of calculations, and the test kappa score ranged between 0.579 and 0.815 (0.0646 SD). As a comparison, the much simpler processing of whole image histograms with a decision tree classifier of depth equal to one yielded a test kappa score of 0.706 +/- 0.0480 SD. This exemplifies the enormous variability of more complex ML models, due to the variability of trainable parameters, and difficulty of obtaining a sufficient number of independent data sets as a consequence of the well documented “data greediness” ^21^ of machine learning (see Figure 7B). This variability may hamper the value of the so-called “grid search” consisting of systematically testing a large number of parameter combinations. This is the reason why we chose to use a limited number of models (as selected in our preliminary study) and a single parameter (cohen kappa score) to assess the success of a given method. This conclusion may seem trivial since is matches the accepted guidelines of medical trials, including as an essential rule the need for a clear definition of tested models and addressed questions, in addition to the use of a sufficiently extensive dataset.

Fourth, despite aforementioned limitations, our study provided an illustration of general guidelines that might facilitate the choice of a strategy for task-specific model development:

i) Attempts at classifying serum samples on the basis of whole field images were fairly unsucessful (Figure 3). Simpler models displayed both insufficient capacity to fit training datasets and high overfitting. Interestingly, convolutional neural networks, that are endowed with high versatility and high capacity to process 2-dimensional images sometimes, yielded a kappa value on the order of 0.6 to classify test datasets. Unfortunately, this value displayed very high variations and incapacity to classify fully unrelated samples (Figure 4). The simplest interpretation might be that the number of samples (1114) was too low as compared to the number of features (i.e the number of pixels that was about one thousandfold higher). Indeed, recently reported successes of ML in processing radiographic images involved training datasets with hundreds of thousands of samples ^51^ ^52^. As an impressive example, a recently reported whole-slide foundation model for digital pathology was trained with more than one billion images ^53^.
ii) Interestingly, as shown on Figure 8, aforementioned models displayed a reasonable capacity to classify 36,163 individual cell images (2,500 pixels each). This emphasizes the key importance of the ratio between sample number and feature number. Note however that this ratio is not the sole determinant of classification efficiency: as shown on Figure 9, combining image segmentation and voting significantly increased classification efficiency in all displayed examples.
iii) Replacing images with histograms resulted in a dramatic performance increase, and several different models yielded kappa values ranging between 0.65 and 0.7 (Figure 5). This supports the reasonable hypothesis that histograms retained much information relevant to positive/negative discrimination, while features related to the geometrical properties of fluorescent zones were clearly lost. Indeed, the number of features per sample was reduced to 256.
iv) Interestingly, our results illustrated the (well known) capacity of ML classifiers to integrate different properties of studied samples. Indeed, in addition to histograms, that contained essentially quantitative information, most models could detect geometric clues, as displayed on Figure 14-A. In addition, the effect of this clue on the training of a convolutional model, as displayed on Figure 14-B, may give us some insight into the training process: a ML model may be viewed as seeking and testing a wide variety of clues, that may be relevant either to specific features of the training data set or features relevant to the intended purpose of training. Figure 14-B suggests that the geometric clue we added was detected within a few epochs, resulting in fairly regular increase of the test-set classification efficiency in contrast to random variations revealing the failure of a model to grasp the expected discrimination property. Thus, between epochs 10 and 26, the standard deviation of test kappa displayed a nearly sevenfold decrease, from 0.0725 to 0.0098.

Fifth, our results are fully consistent with the general concept that optimal classification efficiency and knowledge may be obtained by combining ML and human theoretically-inspired thinking ^54^ ^55^. Indeed, the effect of replacing images with histograms illustrates the capacity of theoretical thinking to improve ML efficiency. Conversely, it must be emphasized that ML was found to outperform ICARE method, since comparable and even slightly increased test kappa scores were obtained without any recourse to DAPI labeling for image segmentation. In addition, the finding revealed by a low depth decision tree classifier suggested that the level of a single intensity level might be a good predictor of positiveness.

Sixth, in view of the capacity of ML to fulfill fairly complex tasks, it may be surprising that no model was found to perform the seemingly simple task of positive/negative discrimination with nearly perfect efficiency. The simplest explanation of this finding might be that, in addition to the distribution of fluorescence intensity, the shape of fluorescent zones might be subtly different in positive and negative samples, which might account for the need of a minimal experience for human observers to perform diagnosis. It might be hypothesized that more extensive datasets might be needed to train ML models to recognize these features in addition to simpler patterns of fluorescence intensity. An attractive prospect would be to use ML to evidence these features and facilitate microscopist training. In this respect, It may be noticed that we used fairly basic ML models despite the impressive successes that were reported with more sophisticated approaches including innovative architectures as exemplified by transformers ^56^, or combination of standard models with procedures such as transfer learning or adversarial learning ^57^. However, while the interest of complex models was recently emphasized, this was not deemed appropriate in the present study due to the simplicity of the classification problem task we addressed and fairly limited dataset size. Note that the potential interest of so-called small-scale artificial intelligence models was recently emphasized ^58^ as well of the advantage of simpler task-specific modesl ^17^.

### Conclusion and Perspective

First, our results support the feasibility of developing a simple and specific model that might be, in a first step, combined with conventional human intervention with the double aim to increase diagnosis safety ^6^ and take advantage of a progressive increase of dataset volume to train more and more efficient models that might reliably achieve a performance comparable to that achieved by experienced human operators. In this respect, the simplest and most attractive approach suggested by our study would be to use a low-depth decision tree to classify image histograms, with an expected kappa index close to 0.71. This seems sufficient to warrant the incorporation of this procedure into actual medical practice. Indeed, diagnostic tests with much lower efficiency are already in use. As an example, prostate specific antigen (PSA) is routinely used to detect prostate cancer, despite a modest kappa index that was reported to range between 0.56 and 0.6 ^59^

Second, the similarity between the outputs of different classifiers and the consistent upper value of kappa to about 0.7 is a strong incentive to question the validity of using human expertise as a gold standard. Indeed, since the introduction of digital image processing in cell biology, marked limitations of the human eye such as limited quantification ability were emphasized ^60^, and this supported the possibility that human observers might be sometimes outperformed by ML ^61^. It would thus be warranted to subject the evolution of the 1114 tested patients included in this study to careful scrutiny in order to determine the possible significance of discrepancies between human and ML classification. This might be highly informative, and the present study may provide an incentive to launch a clinical study to address this point.

## 4. Methods

The methods used in the present study were selected in an exploratory investigation and were fully described in a previous report ^35^. Here we shall summarize the main points and emphasize specific choices. As explained above, it was important to prevent the pitfall of using the extraordinary versatility of ML tools to fit methods to the explored dataset, which might impair the significance of conclusions.

### 4.1 Patients

This retrospective study was performed on 1114 sera that were sent by clinical departments to the immunology laboratory of Marseilles public hospital (APHM) to detect anti neutrophil cytoplasmic antibodies. Samples were part of the Marseilles Biobank (DC 2012-1704) and the study was approved by the health data committee of APHM.

### 4.2 Microscopic tests

Ethanol-fixed human neutrophil slides (Immuno Concepts, Sacramento, Ca, USA) were first incubated with patients’ sera to bind antibodies, then labeled with fluorescein isothiocyanate (FITC)-conjugated anti-human immunoglobulin (Immuno Concept) to reveal bound antibodies and 4,6-diaminophenylindol (DAPI)-containing medium (Vectashield) that was found to stain deposited cells ^35^.

For each patient, two images of the same microscopic field were automatically captured with a 20x objective using excitation wavelengths of 480 nm to reveal FITC and 360 nm to reveal DAPI. This was performed with a robotized fluorescence microscope (Axio Imager M2, Carl Zeiss, Jena, Germany) equipped with an automatic slide handling system (SlideExpress, Märzhäuser, Wetzlar, Germany). Images of 1360×1024 pixel resolution and 256 intensity levels were generated with a CCD camera (ProgRes®MF Cool camera, Jenoptik, Germany). Pixel size was 0.22 µm. Images were examined by an experienced biologist: 217 out of 1114 samples were classified as positive.

### 4.3 Image preprocessing

#### 4.3.1 Segmentation

A custom-made algorithm was used as a plugin for Image J software to perform single cell image segmentation. The basic principle consisted of following the contour of individual cells with a threshold of 60/256 that was found suitable for all 1114 whole field images. These images were then transformed with homothety to fill 50×50 pixel images. A total of 36163 individual images were generated by processing whole field ones.

#### 4.3.2 Other preprocessing operations

Images were converted to numpy arrays in order to perform all further operations with python programming language. In some cases (as indicated in the results section) the following procedures were used:

- Whole field or cell image were converted into histograms, i.e. the frequency distribution of all 256 intensity levels, with numpy histogram function (using as parameters: range=(0,255),bins=256)

- In some cases, intensity normalization was performed with standard scaler or robust scaler provided by scikit-learn (http://scikit-learn.org, version 1.7.0 was used). However, while normalization is an usual step of image processing, it was not always found useful to achieve optimal positive/negative discrimination.

- dimensional reduction was performed with principal component analysis, using scikit-learn PCA function with n_components parameter set at 10. This choice was based on preliminary tests performed in our exploratory investigation.

### 4.4 ML algorithms

We used 7 classifiers of growing complexity. All classifiers were implemented with scikit-learn excepted convolutional networks that were implemented with version 2.15.0 of tensorflow (https://www.tensorflow.org). In all cases, we used default hyerparameters unless otherwise mentioned.

- *K nearest neighbor classifier* (Knn) is arguably the most straightforward classifier that consists of classifying any sample as the majority of its k neighbors in the feature multidimensional space. It must be noticed that the choice is dependent on the definition of distance and this is altered by common preprocessing operations such as normalization (or scaling).
- *Decision trees* (Dt) will partition a dataset with a sequence of binary tests. The number of sequences (hyper parameter max_depth) that is not limited in absence of a specific instruction, was set between 1 and 8, as explained in the result section. Note that decision tree classification is not expected to depend on scaling.
- *Support vector machines* (*Svm*) separate the feature space in domains with fairly complex algorithms^62,63^
- *Random forest classifier* (Rf) and *Gradient boosting classifier* (GB) may be viewed as combinations of decision trees. The principle is clearly explained in ^63^. A decrease of interpretability is the price to be paid for the performance increase.
- *Multilayer perceptron* (Mlp) is a dense neural network. The hidden_layer_sizes parameter was used as (30,30,30), corresponding to three layers of 30 nodes each. This is fairly different from the default value of (100) and it was selected on the basis of several exploratory investigations performed on different models ^64^.
- *Convolutional neural networks* (Cnn) were found to display excellent performances in image analysis, due to their capacity to identify remarkable patterns fairly independently from their localization. We mainly used 2-layer models that were inspired from ^30^ and involved about 1,400,000 parameters. Models involving 1 or 3 layers were also tested.

Compilation was performed with the following parameters:
optimizer=“rmsprop”, loss=“binary_crossentropy”, metrics=[“accuracy”,“AUC”, Kappa()] where Kappa() is a custom made metric written in python.

### 4.5 Defining the quality of ML models

The elaboration of a ML model may be viewed as the selection of the “best” algorithm and parameters. Thus, the metric used to define the so-called best combination is an essential determinant of the final choice. All aforementioned models are endowed with a built-in metric that is used to optimize parameters during training (this is the aforementioned loss parameter). However,Cohen kappa index *k* was used to quantify the success of final models, as indicated in the results section. This parameter is defined as:

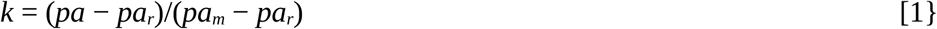

where *k* is kappa index, *pa* is the probability that a given sample be similarly classified by the compared classifiers, *pa_m_* is the index obtained with two identical classifiers and is expected to be equal to 1, and *pa_r_* is the probability of similarity between two random classifications sharing the repartition of samples between classes (i.e. fraction of positive or negative ones).

In each tested combination, unless otherwise mentioned, datasets were randomly divided five times into a training set (75%) and a test set (25%). The training set was used to train the chosen model and this was used to classify the test set. In each case, kappa was calculated with cohen_kappa_score function provided by scikit-learn or with a custom-made python function. Mean values and standard deviation are shown in the results section.

## Author Contributions

D.B., P.B. and N.B. devised the study. D.B. and N.B. managed biological investigations. P.B. performed IA calculations and drafted the manuscript. D.B., P.B. and N.B. elaborated the final version of the manuscript.

## Informed Consent Statement

Informed consent was obtained from all subjects involved in the study.

## Data Availability Statement

All data used in the present study will be communicated on reasonable request to the authors after proper agreement of the Institutional Ethical Board.

## Acknowledgment

All specific apparatus used in this study was funded by the grant “AORC Junior 2017” from the Assistance Publique Hôpitaux de Marseille (APHM, Marseillle France).

## Abbreviations

ANCA: anti-neutrophil cytoplasmic antibodies.
ANA: anti-nuclear antibodies
APHM: Assistance Publique - Hôpitaux de Marseille
Cnn: convolutional neural network
Dt: decision tree classifier
DAPI: 4,6 diaminophenylindole.
FITC: Fluorescein isothiocyanate
Gb: gradient boosting classifier
Knn: kneighbors classifier
ML: machine learning
MLP: Multilayer perceptron classifier (a dense neural network)
Rf: random forest classifier
Svm: support vector machine

